# A machine learning based aging measure among middle-aged and older Chinese adults: the China Health and Retirement Longitudinal Study

**DOI:** 10.1101/2021.04.16.21255644

**Authors:** Xinqi Cao, Guanglai Yang, Xurui Jin, Liu He, Xueqin Li, Zhoutao Zheng, Zuyun Liu, Chenkai Wu

## Abstract

**Background:** Biological age (BA) has been accepted as a more accurate proxy of aging than chronological age (CA). This study aimed to use machine learning (ML) algorithms to estimate BA in the Chinese population.

**Methods:** We used data from 9,771 middle-aged and older (≥ 45 years) Chinese adults in the China Health and Retirement Longitudinal Study. We used several ML algorithms (e.g., Gradient Boosting Regressor, Random Forest, CatBoost Regressor, and Support Vector Machine) to develop new measures of biological aging (ML-BAs) based on physiological biomarkers. R-squared value and mean absolute error (MAE) were used to determine the optimal performance of these ML-BAs. We used logistic regression models to examine the associations of the best ML-BA and a conventional aging measure – Klemera and Doubal method-biological age (KDM-BA) we previously developed – with physical disability and mortality, respectively.

**Results:** The Gradient Boosting Regression model performed best, resulting in a ML-BA with R-squared value of 0.270 and MAE of 6.519. This ML-BA was significantly associated with disability in basic activities of daily living, instrumental activities of daily living, lower extremity mobility, and upper extremity mobility, and mortality, with odds ratios ranging from 1% to 7% (per one-year increment in ML-BA, all P <0.001), independent of CA. These associations were generally comparable to that of KDM-BA.

**Conclusion:** This study provides a valid ML-based measure of biological aging for middle-aged and older Chinese adults. These findings support the application of ML in geroscience research and help facilitate the understanding of the aging process.

## Introduction

Aging is a complex, inevitable, and multifactorial process, characterized by functional deterioration, physiological damage, and multiple age-related diseases [1]. One key question to address aging-related issues is how to accurately and precisely quantify aging. With accumulating evidence supporting the utility of biological age (BA) in predicting age-related outcomes and differentiating individual health status [2-5], BA has therefore been accepted as a more accurate proxy of aging than chronological age (CA). BA is generally referred to a single latent variable that integrated multiple biomarkers relevant to health [6]. Various statistical methods have been used to approximate BA, such as the multiple linear regression method [7], the principal component analysis [8], the Hochschild’s method [9], and the Klemera and Doubal method (KDM) [10]. KDM has been suggested as the optimal method for BA estimation [11]. Machine learning (ML) offers tremendous opportunities for researchers to make predictions from multidimensional data and improve performance in predicting outcomes [1, 12]. However, the application of ML in developing aging measures is limited [2, 13-16]. Most of them were conducted among adults in Europe and the US [13-15], and ML seems not to provide more accurate aging measures than conventional methods [2].

China is facing rapid population aging, which brings formidable challenges to policymakers and caregivers. In 2019, the Chinese population accounted for 18% of the world population, with 164.5 million adults aged 65 and over and 26 million adults aged 80 and over [17]. Developing aging measures for the Chinese population is of great significance to solve aging-related issues in this country, such as facilitating the early identification of adults at risk. To date, a few relevant studies have been conducted in the Chinese population [4, 8, 18-22]. Most of them used the multiple linear regression method [18, 22] or the principal component analyses method [8, 19, 21]. We have previously developed a valid physiological biomarkers-based aging measure using KDM (hereafter referred to as KDM-BA) [4]. It remains unclear whether a ML-based aging measure could be developed in the general Chinese population and how it behaves relative to the KDM-BA we developed.

Therefore, this study aimed to apply several ML algorithms (e.g., Gradient Boosting Regressor, Random Forest, CatBoost Regressor, and Support Vector Machine) to develop new aging measures (hereafter referred to as ML-BAs). We then examined the associations of the best ML-BA and KDM-BA with physical disability and mortality during the follow-up period. We used data from the China Health and Retirement Longitudinal Study (CHARLS), a nationally representative survey.

## Methods

### Study population

Data were from the CHARLS, a nationally representative longitudinal survey of middle-aged and older adults in China. The details of the study design and comprehensive assessments have been described elsewhere [23]. In brief, the CHARLS used a multistage sampling strategy covering 28 provinces, 150 counties/districts, and 450 villages/urban communities across the country. Adults aged 45 years and older were first recruited in 2011/2012, and completed three follow-up visits biennially up to 2017/2018. Ethical approval for collecting data on human subjects was received from the institutional review board at Peking University. Written informed consent was obtained from all participants. Out of 11,847 participants enrolled in the baseline survey (2011/2012) and provided blood samples, we excluded those aged less than 45 years or older than 85 years (N = 1820), with missing data on covariates (N = 256), leaving the analytic sample of 9,771 adults aged 45-85 years. We then assembled various analytic samples for different outcomes due to missingness on each outcome (**Figure 1**).

**Figure 1.**
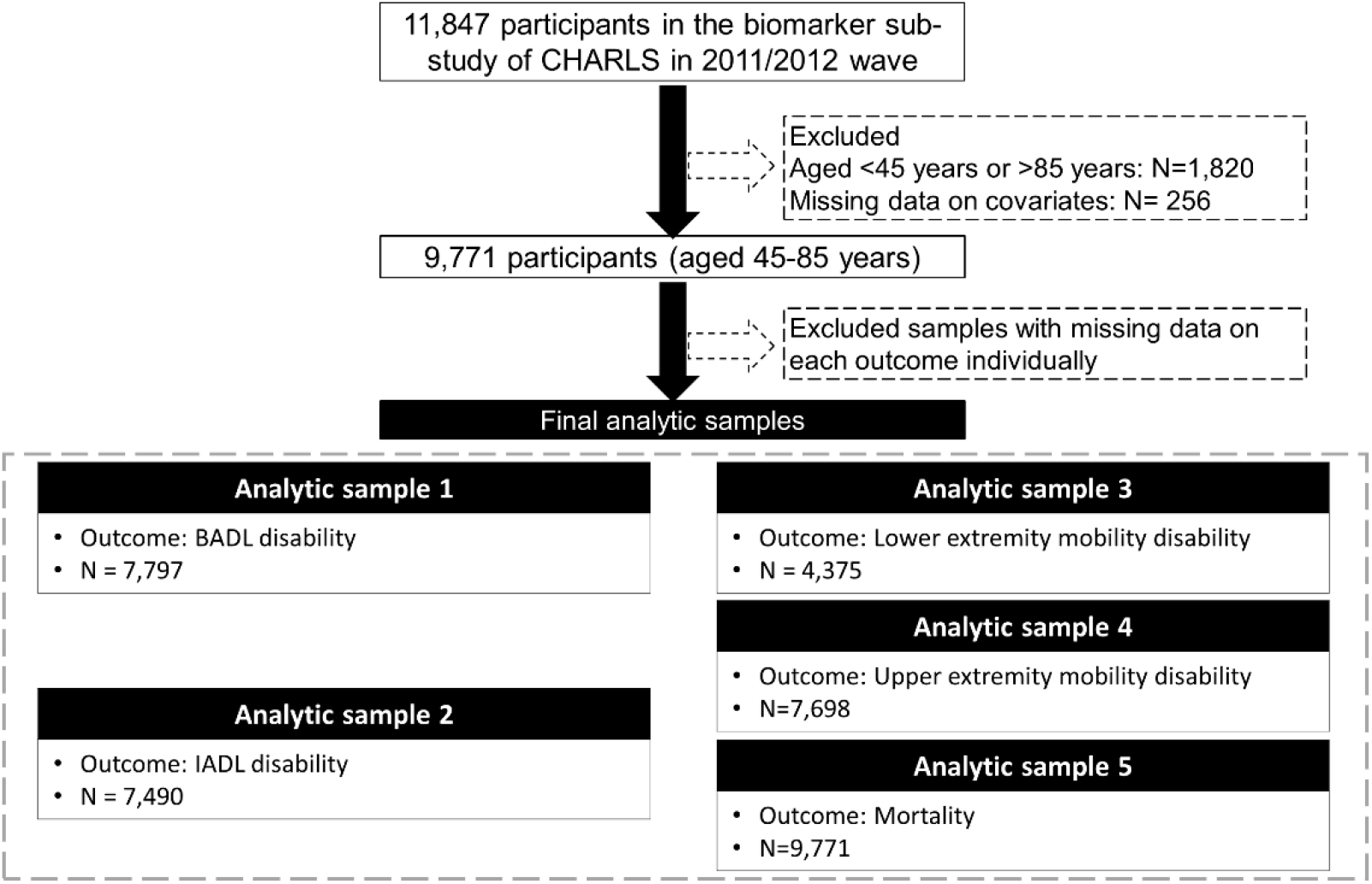
Flow chart of the analytic sample. Note: CHARLS, the China Health and Retirement Longitudinal Study; BADL, basic activities of daily living; IADL, instrumental activities of daily living.

### Biomarker selection and BA calculation

#### ML-BA calculation

Candidate biomarkers were considered based on knowledge about their role in the aging process, application in previous aging studies, and data availability. A total of 16 blood biomarkers (i.e., total cholesterol, triglyceride, glycated hemoglobin, urea, creatinine, high-sensitivity C-reactive protein, platelet count, white blood cell count, mean corpuscular volume, glucose, high-density lipoprotein, low-density lipoprotein, hemoglobin, cystatin, uric acid, and hematocrit) were measured in the 2011/2012 wave of CHARLS [24], plus systolic and diastolic blood pressure, and pulse, resulting in 19 candidate biomarkers for the initial consideration in this study. We firstly imputed the missing data with mean and normalized data using min-max scalar. Then, we trained models with these 19 candidate biomarkers using 10 fold cross-validations to obtain the R-squared value and the mean absolute error (MAE). The models we considered included Gradient Boosting Regressor, Light Gradient Boosting Machine, CatBoost Regressor, Random Forest, Extra Trees Regressor, Support Vector Machine, and AdaBoost Regressor. The final ML-BA in the unit of years was computed.

#### KDM-BA calculation

We used a part of the biomarkers identified above and followed the procedure we previously described [25] to calculate KDM-BA. In brief, the KDM takes information from *m* number of regression lines of CA regressed on *m* biomarkers (*m* = 8 in this study) (i.e., total cholesterol, triglyceride, glycated hemoglobin, urea, creatinine, high-sensitivity C-reactive protein, platelet count, and systolic blood pressure). The final product is KDM-BA in the unit of years.

### Physical disability

The physical function status of basic activities of daily living (BADL) was assessed based on six daily activities, including eating, dressing, transferring, using the toilet, bathing, and continence [26]. Participants were asked if they needed assistance with each of the activities. We categorized participants as having BADL disability if they had incontinence problems or needed assistance in performing at least one of the other five activities (eating, dressing, transferring, toileting, and bathing) [27]. The functional status of instrumental activities of daily living (IADL) was assessed by five instrumental activities, including cleaning the house, managing money, taking medications, shopping for groceries, and preparing the hot meal [28]. We categorized participants as having IADL disability if they needed assistance in performing at least one of the five instrumental activities [27]. Mobility function was divided into the function of upper extremities and lower extremities. The mobility function of the upper extremity was assessed by three activities, including extending arms up, lifting 10 jin, and picking up a small coin. The mobility function of lower extremity was assessed by four activities, including walking 100 m, climbing several flights of stairs, getting up from a chair, and stooping or kneeling or crouching. We categorized participants as having mobility disability if they needed assistance in performing at least one activity [29]. The functional status was assessed at baseline, 2013 wave, and 2015 wave. Since the time of developing disability during the follow-up period was not available, we constructed a binary outcome to denote the occurrence of disability within the 4-year follow-up since baseline.

### Mortality

In CHARLS, the death information was collected from the exit interviews in 2013, 2015, and 2018 waves. However, in the 2015 and 2018 waves, the exact date of death was not available. Therefore, in this study, we constructed a binary variable to denote the occurrence of death within the 6-year follow-up since baseline.

### Covariates

All covariates were obtained at baseline. The sociodemographic variables including age, sex, educational level, marital status, and residence were collected from the self-reported questionnaire. Educational level was defined as no school, primary school, middle school, and high school or above. Marital status was defined as currently married, and others (e.g., separated, divorced, widowed). The residence was defined as urban and rural. Health behaviors including smoking, alcohol drinking, and body mass index (BMI, kg/m^2^) were collected by the structured home interview. Smoking status was defined as current smoker and non-smoker. Alcohol drinking status was defined as current drinker and non-drinker. BMI was calculated as weight in kilograms divided by height in meters squared. We categorized participants as underweight (BMI < 18.5 kg/m^2^), normal (18.5 ≤ BMI < 24.0 kg/m^2^), overweight (24.0 ≤ BMI < 28.0 kg/m^2^), and obese (BMI ≥ 28 kg/m^2^). Disease count was determined by ten self-reported chronic diseases, including hypertension, diabetes or high blood sugar, cancer or malignant tumor, chronic lung disease, heart problems, stroke, kidney disease, stomach or other digestive diseases, arthritis or rheumatism, and asthma. We then divided participants into four groups—no disease, one disease, two diseases, and three or more diseases.

### Statistical analyses

We used 10-fold cross-validations to train ML-BA with 90% training data set and validate it with 10% testing data set. We compared different ML algorithms based on the R-squared value and the MAE. Finally, we selected the Gradient Boosting Regression model to compute the best ML-BA in the unit of years in the total population.

Baseline characteristics of the study population were presented as means ± standard deviations (SD) for continuous variables or numbers (percentages) for categorical variables. To examine the associations of the two aging measures (i.e., ML-BA and KDM-BA) with 4-year physical disability incidence and 6-year mortality risk, we used logistic regression models. Odds ratios (ORs) and corresponding 95% confidence intervals (CIs) were documented. Two models were used in our study. Model 1 was a crude model, whereas model 2 was adjusted for CA.

All statistical analyses were performed using SAS version 9.4 (SAS Institute, Cary, NC), Stata version 15 (Stata Corp, College Station, Texas, USA), and Python version 3.8.3. A P value of < 0.05 (two-tailed) was considered statistically significant.

## Results

The basic characteristics of the study population are presented in **Table 1**. The mean CA of the study population was 59.1 (SD = 9.2) years. Of the 9771 middle-aged and older adults, approximately 44.6% were aged ≥ 60 years, 53.5% were females. The mean CAs of males and females were 59.8 (SD = 9.1) years and 58.5 (SD = 9.2) years, respectively.

**Table 1.** Baseline characteristics of the study population.

|  | **Total**<br>(N = 9,771) | **Male**<br>(N = 4,545) | **Female**<br>(N = 5,226) |
| --- | --- | --- | --- |
| Age, years | 59.1±9.2 | 59.8±9.1 | 58.5±9.2 |
| <60 years | 5,414 (55.4) | 2,361 (52.0) | 3,053 (58.4) |
| ≥60 years | 4,357 (44.6) | 2,184 (48.1) | 2,173 (41.6) |
| ML-BA | 59.4 (5.8) | 60.0 (5.8) | 58.8 (5.8) |
| KDM-BA | 57.0 (9.9) | 58.2 (9.4) | 56.1 (10.3) |
| Sex, female | 5,226 (53.5) | — | — |
| Residence, rural | 6,366 (65.2) | 3,005 (66.1) | 3,361 (64.3) |
| Education |  |  |  |
| No schooling | 2,882 (29.5) | 601 (13.2) | 2,281 (43.7) |
| Primary school | 4,018 (41.1) | 2,182 (48.0) | 1,836 (35.1) |
| Middle school | 1,923 (19.7) | 1,160 (25.5) | 763 (14.6) |
| High school or more | 948 (9.7) | 602 (13.3) | 346 (6.6) |
| Marital status |  |  |  |
| Currently married | 8,156 (83.5) | 3,984 (87.7) | 4,172 (79.8) |
| Others | 1,615 (16.5) | 561 (12.3) | 1,054 (20.2) |
| Smoking status |  |  |  |
| Nonsmoker | 6,797 (69.6) | 1,897 (41.7) | 4,900 (93.8) |
| Smoker | 2,974 (30.4) | 2,648 (58.3) | 326 (6.24) |
| Alcohol consumption |  |  |  |
| Nondrinker | 5,973 (61.1) | 1,522 (33.5) | 4,451 (85.2) |
| Drinker | 3,798 (38.9) | 3,023 (66.5) | 775 (14.8) |
| BMI (kg/m<sup>2</sup>) | 23.5±3.9 | 23.0±3.6 | 24.0±4.1 |
| BMI category * |  |  |  |
| Underweight | 650 (6.8) | 315 (7.1) | 335 (6.5) |
| Normal | 4,990 (52.0) | 2,627 (58.8) | 2,363 (46.1) |
| Overweight | 2,828 (29.5) | 1,149 (25.7) | 1,679 (32.7) |
| Obese | 1,130 (11.8) | 377 (8.4) | 753 (14.7) |
| Disease counts |  |  |  |
| 0 | 2,938 (30.1) | 1,469 (32.3) | 1,469 (28.1) |
| 1 | 3,110 (31.8) | 1,482 (32.6) | 1,628 (31.2) |
| 2 | 2,132 (21.8) | 943 (20.8) | 1,189 (22.8) |
| 3 | 1,591 (16.3) | 651 (14.3) | 940 (18.0) |

Notes: ML-BA, Machine Learning method-biological age; KDM-BA, Klemera and Doubal method-biological age; BMI, body mass index.
\*BMI was calculated as weight in kilograms divided by height in meters squared. Underweight was defined as BMI < 18.5 kg/m<sup>2</sup>; normal was defined as 18.5 ≤ BMI < 24.0 kg/m<sup>2</sup>; overweight was defined as 24.0 ≤ BMI < 28.0 kg/m<sup>2</sup>; and obese was defined as BMI ≥ 28 kg/m<sup>2</sup>.

### Characteristics of ML-BA

We considered Gradient Boosting Regressor, Light Gradient Boosting Machine, CatBoost Regressor, Random Forest, Extra Trees Regressor, Support Vector Machine, and AdaBoost Regressor in our study. R-squared value of models ranged from 0.217 to 0.270, and the MAE of models ranged from 6.619 to 6.877 (**Table S1**). Among them, the Gradient Boosting Regressor model performed best with the highest R-squared value of 0.270 and the lowest MAE of 6.519. Hence, we computed ML-BA using the Gradient Boosting Regression model with 19 biomarkers.

In the total study population, ML-BA ranged from 43 to 82 years, with a mean of 59.4 (SD = 5.8) years. In males, ML-BA ranged from 47 to 82 years, with a mean of 60.0 (SD = 5.8) years. In females, ML-BA ranged from 43 to 81 years, with a mean of 58.8 (SD = 5.8) years. As shown in **Figure 2**, ML-BA and CA were highly correlated (cor = 0.75).

**Figure 2.**
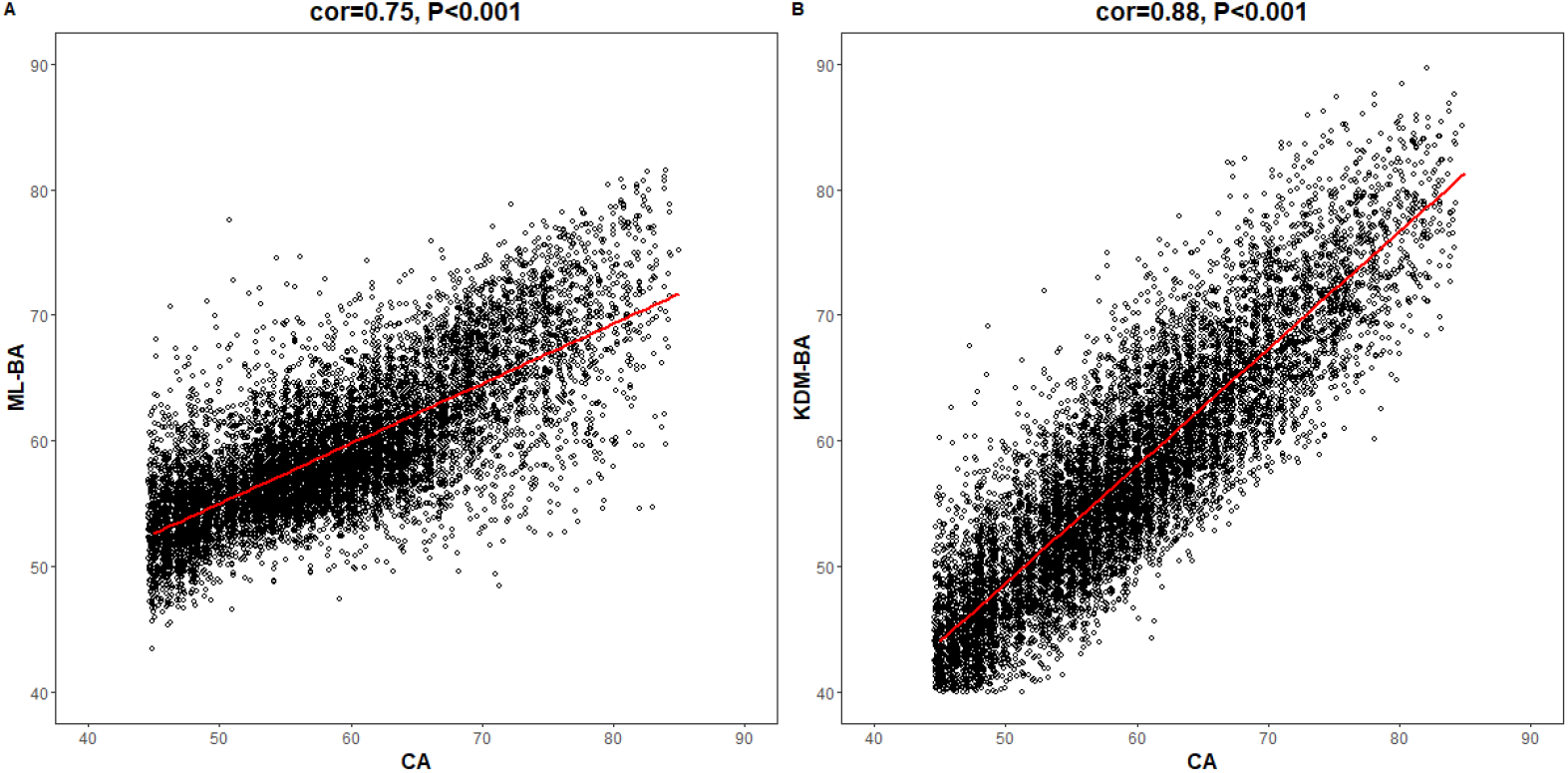
Correlations of CA with ML-BA and KDM-BA. Notes: CA, chronological age; ML-BA, Machine Learning method-biological age; KDM-BA, Klemera and Doubal method-biological age. A and B show the correlation between CA and the 2 measures (ML-BA and KDM-BA), respectively.

### Associations of ML-BA and KDM-BA with physical disability

As shown in **Table 2**, both ML-BA and KDM-BA were significantly associated with 4-year physical disability in the full sample. In the crude model, each one-year increment in ML-BA increased the odds of disability in BADL, IADL, lower extremity mobility, and upper extremity mobility by 6% (OR = 1.06, 95% CI = 1.05, 1.07), 6% (OR = 1.06, 95% CI = 1.05, 1.07), 4% (OR = 1.04, 95% CI = 1.03, 1.05), and 7% (OR = 1.07, 95% CI = 1.06, 1.08), respectively. The strength of these associations was slightly stronger compared to that of KDM-BA. For example, each one-year increment in KDM-BA increased the odds of disability in upper extremity mobility by 4% (OR = 1.04, 95% CI = 1.03, 1.05). Further subgroups analyses by sex did not change the results substantially.

**Table 2.** Unadjusted associations of CA, ML-BA, or KDM-BA with disability and mortality in the full sample and sex subgroup.

|  |  | **BADL disability** | **IADL disability** | **Lower extremity**<br>**mobility disability** | **Upper extremity**<br>**mobility disability** | **Mortality** |
| --- | --- | --- | --- | --- | --- | --- |
| **No. of events/No. of participants** |  | 1,860/7,797 | 1,935/7,490 | 2,380/4,375 | 1,947/7,698 | 882/9,771 |
| **Total** | CA only | 1.048 (1.042, 1.054) | 1.045 (1.039, 1.051) | 1.03 (1.02, 1.04) | 1.05 (1.04, 1.06) | 1.11 (1.10, 1.12) |
|  | ML-BA only | 1.06 (1.05, 1.07) | 1.06 (1.05, 1.07) | 1.04 (1.03, 1.05) | 1.07 (1.06, 1.08) | 1.16 (1.14, 1.17) |
|  | KDM_BA only | 1.043 (1.037, 1.048) | 1.037 (1.031, 1.043) | 1.024 (1.017, 1.031) | 1.04 (1.03, 1.05) | 1.104 (1.096, 1.113) |
| **Male** | CA only | 1.06 (1.05, 1.07) | 1.06 (1.05, 1.07) | 1.04 (1.03, 1.05) | 1.05 (1.04, 1.04) | 1.10 (1.09, 1.12) |
|  | ML-BA only | 1.07 (1.06,1.09) | 1.08 (1.07,1.10) | 1.06 (1.05, 1.07) | 1.08 (1.06,1.09) | 1.14 (1.13, 1.16) |
|  | KDM-BA only | 1.06 (1.05, 1.07) | 1.05 (1.04, 1.06) | 1.04 (1.03,1.05) | 1.05 (1.04, 1.06) | 1.10 (1.09, 1.11) |
| **Female** | CA only | 1.046 (1.038, 1.054) | 1.043 (1.035, 1.051) | 1.03 (1.02,1.04) | 1.055 (1.047, 1.064) | 1.13 (1.11, 1.14) |
|  | ML-BA only | 1.06 (1.05, 1.08) | 1.06 (1.05, 1.07) | 1.04 (1.02,1.05) | 1.08 (1.06, 1.09) | 1.17 (1.15, 1.19) |
|  | KDM-BA only | 1.04 (1.03, 1.05) | 1.036 (1.029, 1.044) | 1.03 (1.02, 1.04) | 1.045 (1.037, 1.052) | 1.11 (1.10, 1.12) |

Notes: CA, chronological age; ML-BA, Machine Learning method-biological age; KDM-BA, Klemmera and Doubal method-biological age; BADL, basic activities of daily living; IADL, instrumental activities of daily living.
Participants with prevalent disability in BADL/IADL/lower extremity mobility/upper extremity mobility were excluded for analyses of BADL/IADL/lower extremity mobility/upper extremity mobility, respectively.

**Table 3** shows the associations of ML-BA and KDM-BA with physical disability when adjusting for CA in the full sample. ML-BA was still significantly associated with all functional disabilities, with ORs ranging from 1.01 to 1.02. Significant association of KDM-BA with disability in BADL was observed, with OR of 1.01 (95 % CI = 1.00, 1.03).

**Table 3.** Risk estimates of disability and mortality predicted by ML-BA and KDM-BA adjusting for CA.

| **Model** | **Variable** | **BADL disability** | **IADL disability** | **Lower extremity mobility disability** | **Upper extremity mobility disability** | **Mortality** |
| --- | --- | --- | --- | --- | --- | --- |
| **No. of events/No. of participants** |  | 1,860/7,797 | 1,935/7,490 | 2,380/4,375 | 1,947/7,698 | 882/9,771 |
| **CA+ ML-BA** | CA | 1.04 (1.03, 1.05) | 1.04 (1.03, 1.05) | 1.02 (1.01, 1.03) | 1.04 (1.03, 1.05) | 1.08 (1.06, 1.09) |
|  | ML-BA | 1.01 (1.00, 1.03) | 1.02 (1.00, 1.03) | 1.02 (1.00, 1.03) | 1.02 (1.01, 1.03) | 1.07 (1.05, 1.09) |
| **CA+ KDM-BA** | CA | 1.04 (1.02, 1.05) | 1.04 (1.03, 1.06) | 1.03 (1.01, 1.04) | 1.05 (1.03, 1.06) | 1.06 (1.05, 1.08) |
|  | KDM-BA | 1.01 (1.00, 1.03) | 1.00 (0.99, 1.01) | 1.00 (0.99, 1.02) | 1.00 (0.99, 1.01) | 1.05 (1.04, 1.07) |

Notes: CA, chronological age; ML-BA, machine learning method-biological age; KDM-BA, Klemra and Doubal method-biological age; BADL, basic activities of daily living; IADL, instrumental activities of daily living.
Participants with prevalent disability in BADL/IADL/lower extremity mobility/upper extremity mobility were excluded for analyses of BADL/IADL/lower extremity mobility/upper extremity mobility, respectively.

### Associations of ML-BA and KDM-BA with mortality

**Table 2** presents the associations of ML-BA and KDM-BA with 6-year mortality in full sample and subgroups by sex. Both ML-BA and KDM-BA were positively associated with mortality risk. The results of the association between KDM-BA and mortality were previously reported [4]. In full sample, each one-year increment in ML-BA and KDM-BA increased the risk of mortality risk by 16% (OR = 1.16, 95% CI = 1.14, 1.17) and 10% (OR = 1.104, 95% CI = 1.096, 1.113), respectively. When stratified by sex, the ORs of ML-BA for mortality risk ranged from 1.14 to 1.17, consistent with that in the full sample (OR = 1.16). Similar results were found for KDM-BA.

After adjusting for CA, both ML-BA and KDM-BA were significantly associated with 6-year mortality risk, although the strength of the associations was attenuated. Each one-year increment in ML-BA and KDM-BA increased the risk of mortality by 7% (OR = 1.07, 95% CI = 1.05, 1.09) and 5% (OR = 1.05, 95% CI = 1.04, 1.07), respectively (**Table 3**). The results suggested that they capture something above and beyond what can be explained by CA alone when predicting mortality.

## Discussion

In this study, we successfully developed an aging measure using the Gradient Boosting Regression model in a sample of middle-aged and older Chinese adults. We found that this ML-BA was predictive of physical disability and mortality during the follow-up period, and these associations were independent of CA. The results were comparable to that of KDM-BA, supporting the development of ML-BA. This ML-BA may serve as a proxy of life span in gerontology and help with the risk stratification in the general Chinese older adults.

To date, several studies have shown that BA calculated using ML has the predictive ability for mortality risk in populations of different countries, such as the US [14, 16], Italy [30], and Singapore [2]. Because of differences in genetic and socio-environmental factors, the findings may not be generalized to various populations in other countries, such as the Chinese population, a rapidly increasing segment worldwide. To the best of our knowledge, no studies have been performed to develop BAs using ML and evaluate the associations of ML-BAs with adverse outcomes in the Chinese population. We filled up this gap in this study. More importantly, we demonstrated that the best ML-BA performed just as well as KDM-BA, which has been suggested as the best biological aging measure [25]. The findings support that ML could be used to develop measures of biological aging. Moreover, both ML-BA and KDM-BA could be developed across various populations separately, and they may capture something underlying the aging process.

It should be noted that the strength of the associations between ML-BA and physical disability, as well as mortality, is slightly stronger than KDM-BA. ML-BA in our study was computed based on 19 biomarkers, while KDM-BA was computed based on only 8 of the 19 biomarkers. The remaining 11 biomarkers included diastolic blood pressure, pulse, white blood cell count, mean corpuscular volume, glucose, high-density lipoprotein, low-density lipoprotein, hemoglobin, cystatin, uric acid, and hematocrit, which had been demonstrated to be associated with aging [31-35]. Hence, we assume that the better performance of ML-BA may be due to the more information covered by ML-BA than that by KMD-BA. ML-BA was developed without prior assumptions and was not dependent on intermediate results from multiple linear regression models [36], making ML-BA easy to be verified. In general, ML-BA may therefore provide a useful tool to identify individual risk for adverse outcomes. Future detailed explorations of ML-BA are needed to facilitate the understanding of aging processes.

The stable associations of ML-BA and KDM-BA with physical disability and mortality risk can be interpreted by the biological biomarkers used to develop the two aging measures. The aging process is subclinical, characterized by various types of biological degradation. So, it is proposed to evaluate aging based on cellular and molecular hallmarks [37]. In our study, biomarkers used for ML-BA and KDM-BA computation were selected from various candidate biomarkers, representing different domains of physiological function: immune system (e.g., high-sensitivity C-reactive protein, platelet count, and white blood cell), cardiac-metabolic system (e.g., Total cholesterol, systolic blood pressure, and low-density lipoprotein), and kidney system (e.g., urea, creatinine, cystatin, and uric acid). First, the immune system is a homeostatic system that helps to maintain the function of the organism, and age-related change in immune function has been demonstrated to affect longevity [38]. Due to infectious disease, older adults usually have an increased risk of morbidity and mortality [39], emphasizing the importance of maintaining the function of the immune system during the aging process. Second, since the incidence of heart disease increases sharply with age, it has been postulated that aging and cardiovascular disease are interrelated [40] and may share common pathology [41]. During the normal aging process, the cardiac-metabolic function is impaired with the increase of age [40], which might contribute to adverse age-related outcomes. Finally, evidence has suggested that even in the absence of comorbidities, the kidney may experience significant age-related changes in structure and function [42]. This implies that the deterioration of kidney function may be one of the important phenotypes of the aging process. The aging measures we developed in the current study integrated various biomarkers of immune function, cardiac-metabolic function, and kidney function; therefore they could evaluate the aging process through multiple physiological systems and work well in predicting physical disability and mortality.

In this study, the large sample size and the nationwide prospective cohort study provide us with the opportunity to develop aging measures by ML and explore its associations with adverse health outcomes in middle-aged and older Chinese adults. Nevertheless, limitations in this study should be acknowledged. First, the relatively short follow-up period (i.e, up to 6 years) of the CHARLS has impeded us to explore the long-term effect of aging measures on the outcomes. Longitudinal studies with long-term follow-up are needed to confirm the associations. Second, we did not have data on the exact timing of the physical disability and death. Therefore, we cannot evaluate the impact of BAs on survival time. Finally, we did not have data on the incidence of chronic diseases (e.g, diabetes, heart disease, and stroke), impeding us to better evaluate the associations of biological aging with chronic diseases.

## Conclusion

In summary, this study provides a valid ML-based measure of biological aging for middle-aged and older Chinese adults. We further demonstrated that this ML-BA was associated with physical disability incidence and mortality. These associations were comparable to that of KDM-BA, a valid physiological biomarkers-based aging measure we have previously developed. The findings support the application of ML in geroscience research and promote further understanding of the aging process. Together with KDM-BA, these aging measures could serve as a proxy of life span and help with the risk stratification in the general Chinese older adults.

## Supporting information

STROBE_checklist

## Data Availability

The CHARLS data are publicly available through the CHARLS website at http://charls.pku.edu.cn/en.

## Author Contributions

ZL and CW conceived and designed the study. XC, GY, and XJ performed the analysis and wrote the initial draft of the manuscript. LH, XL, ZZ, ZL, and CW helped to interpret the results and edit the manuscript. ZL and CW contributed to the critical revision of the manuscript for important intellectual contents. All authors have read and agreed to the final version of the manuscript.

## Data Availability Statement

The the China Health and Retirement Longitudinal Study (CHARLS) data are publicly available through the CHARLS website at http://charls.pku.edu.cn/en.

## Acknowledgments

We thank all respondents of the China Health and Retirement Longitudinal Study (CHARLS).

---

**Table S1.** MAE, MSE, RMSE, and R-squared value of machine learning models.

| **Model** | **MAE** | **MSE** | **RMSE** | **R-squared value** |
| --- | --- | --- | --- | --- |
| Gradient Boosting Regressor | 6.519 | 64.127 | 8.001 | 0.270 |
| Light Gradient Boosting Machine | 6.532 | 64.875 | 8.049 | 0.261 |
| CatBoost Regressor | 6.527 | 65.121 | 8.063 | 0.258 |
| Random Forest | 6.557 | 65.126 | 8.065 | 0.258 |
| Extra Trees Regressor | 6.576 | 65.330 | 8.075 | 0.256 |
| Support Vector Machine | 6.655 | 68.141 | 8.248 | 0.224 |
| AdaBoost Regressor | 6.877 | 68.804 | 8.289 | 0.217 |

Notes: MAE, Mean Absolute Error; MSE, Mean Square Error; RMSE, Relative Mean Square Error.

